# RELATIONSHIP BETWEEN ICU WAITING AND MORTALITY RATE IN PATIENTS UNDER MECHANICAL VENTILATION ADMITTED IN EMERGENCY ROOM

**DOI:** 10.1101/2020.06.03.20121616

**Authors:** Layanna Alves da Silva Andrade, Mônica Lúcia Soares Borges, Geovane Rossone Reis, Aktor Hugo Teixeira, Rebeca Oliveira Crispim da Silva, Izabella Kássia Teixeira dos Santos, Gilvanea Kézia Gomes de Abreu, Amanda Silva de Aguiar

## Abstract

**Background:** The number of patients who remain on prolonged mechanical ventilatory assistance has recently increased. The average length of hospital stay intervenes directly with the number of beds essential for assistance to a given population and is weighted as an indicator of service efficiency.

**Objective:** To investigate the average time in which patients on mechanical ventilation admitted to the emergency department of a hospital remain waiting for a place in the ICU.

**Materials and Methods:** Quantitative field research, where data collection was performed using data from medical records, admission books, death books / outcome of hospitalized patients and in line with pre-established criteria under mechanical ventilation. Mechanical ventilation patients admitted to the ER and the ICU, over 18 years old, were recruited.

**Results:** The research comprises a sample of 67 patients. The average hospital stay was 334 hours and 36 minutes. The general ratio was for every 03 patients who entered for treatment, 01 was discharged and 02 died. The total mortality rate of the sample was 68.65%.

**Conclusion:** The length of stay in ER and ICU in patients on mechanical ventilation is related to the high mortality rate.

## BACKGROUND

According to Christ (2010)^1^; Last (2011)^2^; Becker (2015)^3^ the demand for patients who request emergency services has increased in recent decades, due to the overcrowding of this hospital environment, becoming a worldwide reality. Urgent care according to the Health System has been a priority in Brazil since 2003^4,5^. Pronouncing itself in the National Emergency Care Policy (PNAU). In 2011, the Ministry of Health restructured the urgency policy with new guidelines, suggesting the provision of an urgency and emergency network that includes pre-hospital, hospital and home care components^6^.

Welcoming with Risk Assessment and Classification (AACR), within the scope of emergency care, has become a technology used by the Ministry of Health in the sense of reorienting assistance policy in emergency services, articulating the values of humanization and qualification of assistance^7^.

According to the Ministry of Health (2012)^8^, the risk classification models are located in five levels of paramount importance within the hospital, which are: Level 1 – red room (emergency) which covers immediate risk to life, resuscitation and immediate medical evaluation; Level 2 – orange room (very urgent) which presents imminent risk to life, emergency, medical evaluation within 10 minutes; Level 3 – yellow room (urgent) which shows a potential threat to life / urgency, urgent, medical evaluation within 30 minutes; level 4 – green room (not very urgent) that covers a situation of potential urgency, or of complications, important severity, medical evaluation within 60 minutes; Level 5 – less urgent blue room (non-urgent) or clinical administrative problems, with medical evaluation within 120 minutes.

Within the criteria for the qualification of ICUs (intensive care units) among hospital institutions, it covers the emergency doors with at least 90% of the average monthly occupancy rates. This occupancy rate implies the high rates of patients who require intensive care by the emergency department, directing them to other inappropriate hospital units. Due to overcrowding in the ICUs, patients who need care in this unit are refused, so the amount of death is greater in this group compared to patients admitted to the ICU. However, the demand from the ICUs is capable of expanding their care operations, preserving the quality of care while maintaining the demand for deaths consistent with the severity profile of patients^9^. The average length of hospital stay intervenes directly in the number of beds essential for assistance to a given population and is weighted as an indicator of service efficiency^10^. According to the National Association for Medical Direction of Respiratory Care (NAMDRC), in a 2004 consensus, the number of patients who remain on prolonged mechanical ventilatory assistance has been increasing a lot lately, due to the eventuality where ICU care is more appropriate and technological advancement^11^.

An international prospective study reports that patients who needed Invasive Mechanical Ventilation, had an average stay under ventilatory support of 7 days and an ICU stay of 13 days^12^. This study found that 50% of the patients were not extubated in the first 24 hours, as they spent more than 7 days in the ICU. Similar results were reported by Higgins et al, where the use of IMV was associated with infection and long ICU stay. Thus, it is believed that the use of IMV can worsen the patient’s prognosis and the longer the maintenance period, the longer the patient stays in the ICU^12^.

The ICU has complex technological resources and needs a team that can play a decisive role in the care of critically ill patients, being able to reverse the situation in which they find themselves^13,14^. The procedures are usually performed with a great technological support, specialized professionals and with high-cost subsidies, with the intention of stabilizing patients’ organic dysfunctions and enabling the execution of complex procedures, with this these units represent about 25% of the total financial resources of hospitals^15^. The ICU is the place that centralizes the care of individuals who are serious or at risk of death. They have uninterrupted medical assistance, adding human resources, materials and equipment with access to other technologies for the diagnosis and treatment of patients within the unit^16,17^.

According to Lemos (2005)^18^, predictors of evolution and deaths are widely studied and applied to define the best management of financial resources, modify therapeutic procedures, monitor the performance of the ICU, or compare different units among themselves. The specialized multidisciplinary team is of paramount importance in the care of critically ill patients, however this same team needs constant improvement to maintain excellence in the advances in intensive health care^19^.

On the other hand, emergency services consist of the guidelines of specific legislation, and to the detriment of the systematization of care, resolution and effectiveness of procedures. The emergency is divided into groups, grouping patients according to priority, and the care provided is based on the clinical condition and its respective risks^20,21^. The Resolution No. 7 of February 24, 2010, which provides for the minimum requirements for the operation of Intensive Care Units, it shows us that the infrastructure must contribute to maintaining the patient’s privacy, without interfering with its monitoring; and that they must occupy distinct and exclusive rooms. A medical technician responsible, a nurse coordinator of the nursing team must be formally appointed and a physical therapist coordinating the physiotherapy team, as well as their respective substitutes.

In addition to a daily / routine doctor for each 10 (ten) beds or fraction; a physician on duty at least for every 10 (ten) beds or fraction; Assistance nurses being at least 01 (one) for each 08 (eight) beds or fraction; Physiotherapists, at least 01 (one) for every 10 (ten) beds or fraction; nursing technicians, at least 01 (one) for every 02 (two) beds. These professionals must be available full-time to assist patients admitted to the ICU, during the time they are scheduled to work in the ICU^22^. According to Ordinance No. 895, of March 31, 2017, the ICU is a hospital service for users in serious or at-risk clinical, clinical or surgical situations, requiring intensive care, uninterrupted, with continuous monitoring for 24 (twenty-four) hours.

Therefore, it is necessary to comply with the following Humanization requirements: a) Noise control; b) Lighting control; c) Air conditioning; d) Natural lighting; e) Guarantee of daily and scheduled visits by family members; f) Guarantee to accompany the elderly, in accordance with the provisions of specific legislation; g) Guarantee of information on the evolution of patients to family members, by the medical team at least once a day. Thus, ensuring adequate care for patients^23^. According to Ordinance No. 354, of March 10, 2014, the urgency and emergency service must present a documented organizational structure; preserving the patient’s identity and privacy, ensuring an environment of respect and dignity; promote a welcoming environment and offer guidance to the patient and family in clear language, about the state of health and the assistance to be provided, from admission to discharge.

Every urgent and emergency service must have a technically responsible person with medical training, legally qualified to assume responsibility for 1 (one) urgent and emergency service. In the event of the absence of the technician responsible, the service must have a professional legally qualified to replace him; having a medical team in sufficient quantity to provide care for 24 hours^24^. The physical infrastructure is dimensioned according to the demand, complexity and care profile of the unit, ensuring the safety and continuity of patient care, covering some essential points such as the risk classification room; room to file medical records or patient care records; doctor’s offices; sanitation area; procedure room with area for suturing, recovery, hydration, and medication administration; nebulization area; room for resuscitation and stabilization; observation and isolation rooms; nursing post; drugstore; among others.

As the ASSISTANCE OPERATIONAL PROCESSES of this same ordinance is treated in its subitem 8.3.1 “When the transfer to an Intensive Care Unit is necessary, it must be carried out as soon as possible”. Thus, structural and care differences and in the management of critically ill patients, especially dependent on mechanical ventilation, between admission to an emergency room and an intensive care unit are clear^24^.

The IBGE (2015)^25^, National Register of Health Establishments of Brazil –CNES (2015), there are around 20,804 beds in the intensive care unit (ICU) among Brazilian capitals. For every 10,000 inhabitants there is a proportion of 4.16 ICU beds per person. In the Unified Health System (SUS) there are approximately 8,930 beds, with a variation of 2.07 beds for 10,000 inhabitants. In the private sector, 11,874 ICU beds are determined in an alternation of 5.79 beds for 10,000 inhabitants. Northern Brazil is the region with the least ICU beds, based on a 5% proportion, totaling 2,058 ICU beds. Above all, the southeast region has more than half of the ICU beds in Brazil, about 22,200 beds in the proportion of 54.2% of the country. According to Intensive Care Medicine in Brazil (2015), National Registry of Health Facilities in Brazil – CNES (2015); ministry of health (2015) a survey was conducted in Brazil, between 2010 and 2015, showing that the beds of the intensive care unit (ICU) in general, by SUS and private individuals, had an increase in this period. Qualifying that adult ICU beds in general there was an increase of 24%, infant ICU beds increased by 13% and neonatal ICU beds increased by 16%. Under the Unified Health System (SUS), adult ICU beds increased by 27%, child ICU beds increased by 12% and neonatal ICU beds increased by 23%. In the private sector, the number of beds in the adult ICU was 21%, in children’s ICU beds 13%, and in neonatal ICU beds 9%^26, 27, 28^.

The criteria for admission to the intensive care unit (ICU), intended for hospitalization for these potentially critical or recoverable patient beds, due to the increased cost, insufficient supply and high demand, at the time of the indication the request must be done carefully with specific age groups: Neonatal patients are newborns from 0 to 28 days old, pediatric patients from 29 days to 12 years old, and adult patients older than 13 years29. The prioritization criteria for admission to the ICU are composed of three priority groups where in the first group are critical patients who are in a coma or not, and unstable who need care that only in the ICU can be promoted. The second group is aimed at patients who need intensive monitoring or need immediate intervention. In the third group, they are critical patients, but who have a reduced probability of survival due to the underlying disease or the nature of an acute disease^29^.

In the medical literature, deaths of patients admitted to the ICU are classified in two ways: a) unexpected death (when it happens even after the use of all accessible therapy, for example, in the consecutive period of trauma or septic shock) and b) death expected (occurs in long unsuccessful treatments, as we can relate them to inoperable tumors, chronic diseases and the presence of multiple organ failure)^30^. According to the demand of patients with severe health conditions who need continuous monitoring and intensive care through hospitalization in beds of the Intensive Care Unit (ICU) offered by the Unified Health System (SUS), this study aimed to investigate the average time in which patients on mechanical ventilation admitted to the emergency department of a hospital remain waiting for a place in the ICU.

## MATERIALS AND METHODS

The method for the selection of the quantitative study was sampling performed using data from medical records, admission books, death books / outcome of hospitalized patients and in line with the pre-established criteria under mechanical ventilation, in the sectors of the First Aid adult – ER and in the intensive care unit – ICU of Hospital Regional Público de Gurupi

- HRPG, from September 2019 to January 2020.

The study population in general was 82 patients of both sexes. According to the exclusion criteria, 15 patients were excluded due to 3 minors, 6 patients who were transferred to another hospital, and 6 patients who did not obtain an outcome at the end of the research. In the final result, the research remained with 67 patients.

The theoretical basis of the research was carried out through the databases SciELO, Google Scholar and Lilacs, emphasizing the languages Portuguese, English and Spanish, with the following descriptors Mechanical Ventilation, Emergency, ICU, Hospitalization and Death. Data collection started after approval by the research ethics committee involving human beings at the regional public hospital in Gurupi (CAAE: 19654719.4.0000.5518), in accordance with resolution 466/12 and resolution 510/16 both from the Ministry of Health. Cheers. Visits for data collection were carried out daily, in the morning, afternoon and evening periods. The identification form was conducted through a continuous evaluation of the patients, where they were evaluated from the ER, when they were transferred to the ICU until their final outcome, being for death or discharge from the ER or ICU. Individuals under 18 years of age, who were not admitted to the ER and ICU sectors, who did not depend on mechanical ventilation, and / or were admitted to the hospital under mechanical ventilation, however, with death decree through a death certificate, were excluded.

Regarding the statistical analysis, the correlation data between the quality indicators (length of stay and deaths) were analyzed using Pearson’s and / or Staverman’s coefficient in order to assess the degree of linear association. After validation, they were compared using the Student test paired using the SPSS® software for parametric variables and for nonparametric chi-square with a significance level of 5%. The values were demonstrated by means of tables and graphs generated by STATA®, considering the significance level of p < 0.05 or 5%.

## RESULTS

The research comprises a sample of 67 patients, of which 38.8% (n = 26) are female and 61.2% (n = 41) are male. Patients are between the ages of 18 to 97 years of age, with the age group of 70 to 79 years with the largest representation in the sample, with 35.8% (n = 24) followed by the age group of 80 to 89 years with 22.3% (n = 15). This information is contained in Table 1.

**Table 1.**
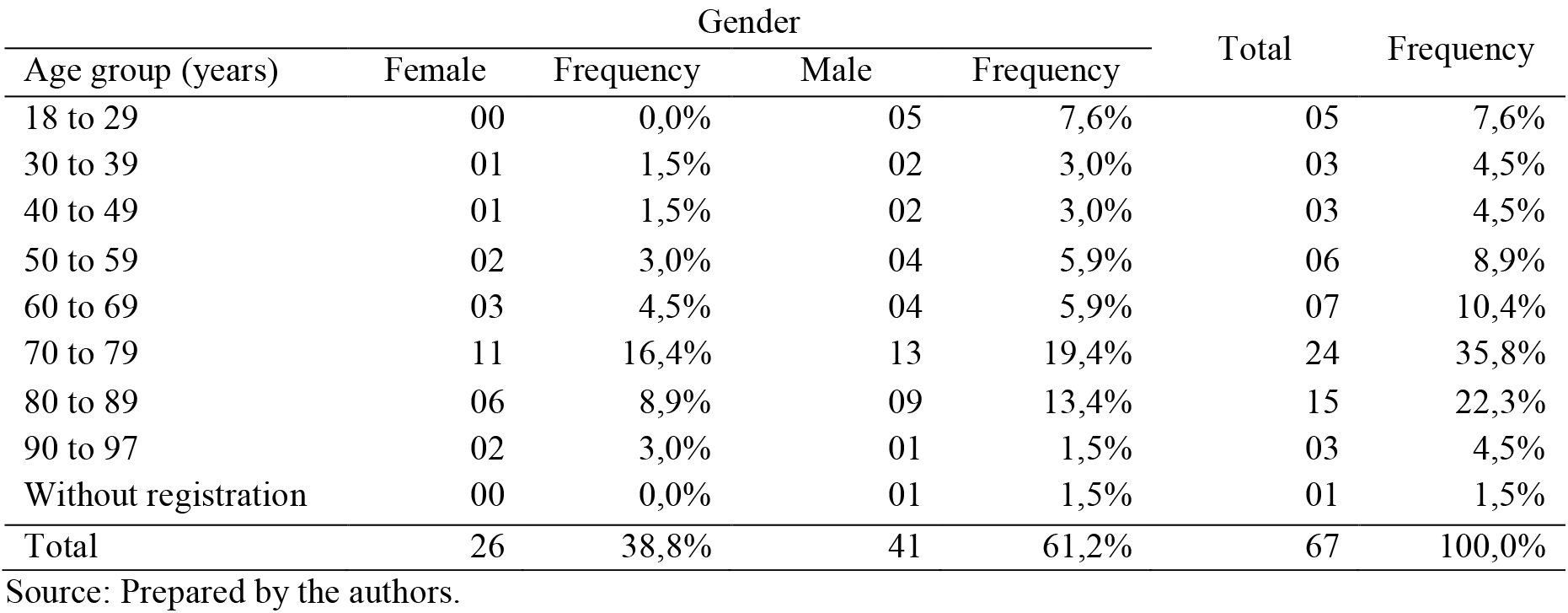
Classification of the sample by age and gender. Gender Total Frequency

| Age group (years) | Gender |  |  |  | Total | Frequency |
| --- | --- | --- | --- | --- | --- | --- |
|  | Female | Frequency | Male | Frequency |  |  |
| 18 to 29 | 00 | 0,0% | 05 | 7,6% | 05 | 7,6% |
| 30 to 39 | 01 | 1,5% | 02 | 3,0% | 03 | 4,5% |
| 40 to 49 | 01 | 1,5% | 02 | 3,0% | 03 | 4,5% |
| 50 to 59 | 02 | 3,0% | 04 | 5,9% | 06 | 8,9% |
| 60 to 69 | 03 | 4,5% | 04 | 5,9% | 07 | 10,4% |
| 70 to 79 | 11 | 16,4% | 13 | 19,4% | 24 | 35,8% |
| 80 to 89 | 06 | 8,9% | 09 | 13,4% | 15 | 22,3% |
| 90 to 97 | 02 | 3,0% | 01 | 1,5% | 03 | 4,5% |
| Without registration | 00 | 0,0% | 01 | 1,5% | 01 | 1,5% |
| Total | 26 | 38,8% | 41 | 61,2% | 67 | 100,0% |

Source: Prepared by the authors.

The patients who made up the sample had their treatment monitored between September 4, 2019 to 07 and January 2020. All patients were admitted to the ER and underwent the intubation process. Of the total, 45 patients were admitted to the ICU and of these 23 patients were extubated. In this sample, there were no cases of patients transferred for treatment outside the home. Table 2 shows the mortality rates and the average length of stay in all treatments, in ER, in the ICU and with the use of MV.

**Table 2.** Mortality Rate and Average Length of Hospitalization

| All treatments |  |  |  |  |  |
| --- | --- | --- | --- | --- | --- |
| Sample | Death | Discharge | In treatment | Mortality Rate | Average time |
| 67 patients | 46 | 20 | 01 | 68,65% | 334:36h |
| Treatment in ER |  |  |  |  |  |
| Sample | Death | Discharge | In treatment | Mortality Rate | Average time |
| 67 patients | 16 | 06 | 45 | 23,88% | 161:52h |
| ICU treatment |  |  |  |  |  |
| Sample | Death | Discharge | In treatment | Mortality Rate | Average time |
| 45 patients | 30 | 14 | 01 | 66,67% | 275:14h |
| Mechanical ventilation treatment |  |  |  |  |  |
| Sample | Death | Discharge | In treatment | Mortality Rate | Average time |
| 23 patients | 5 | 18 | 00 | 21,74% | 252:15h |

Source: Prepared by the authors.

**Graph 1.**
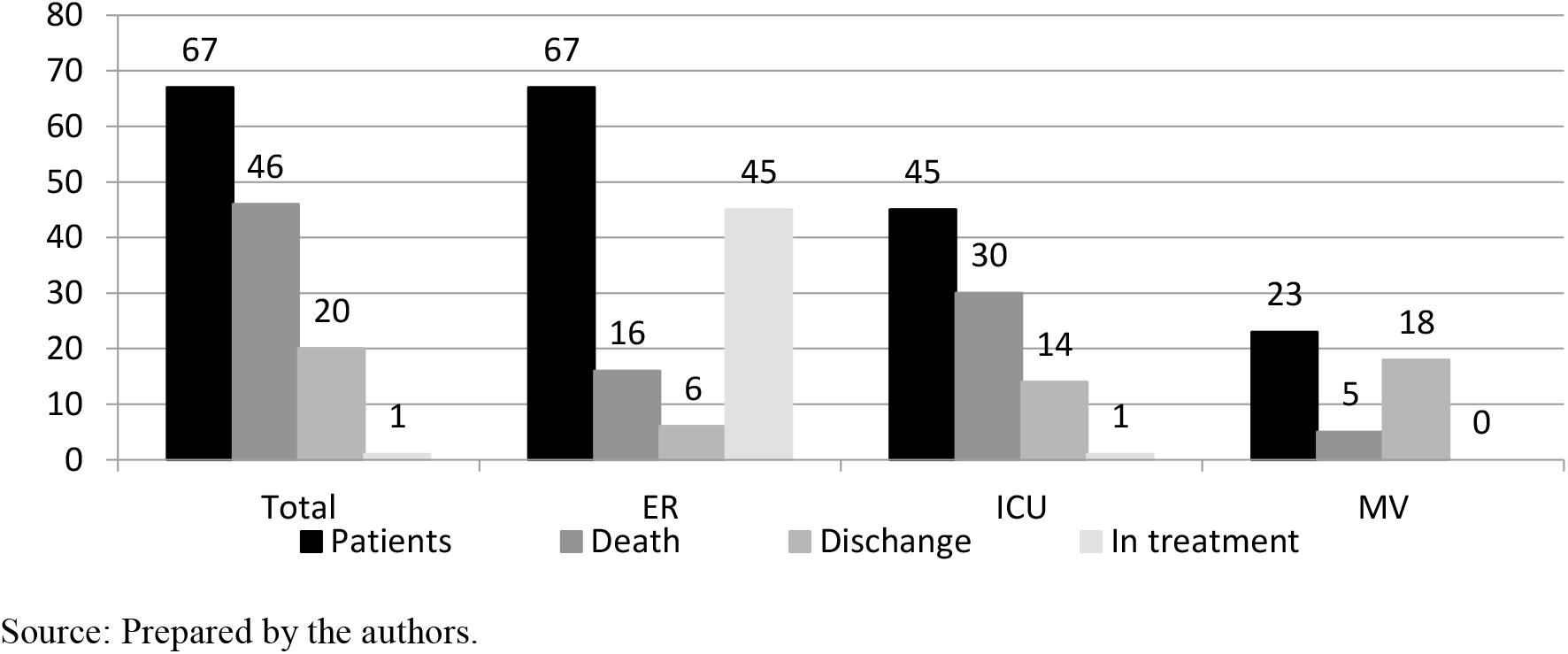
Description of the sample by type of treatment

**Graph 2.**
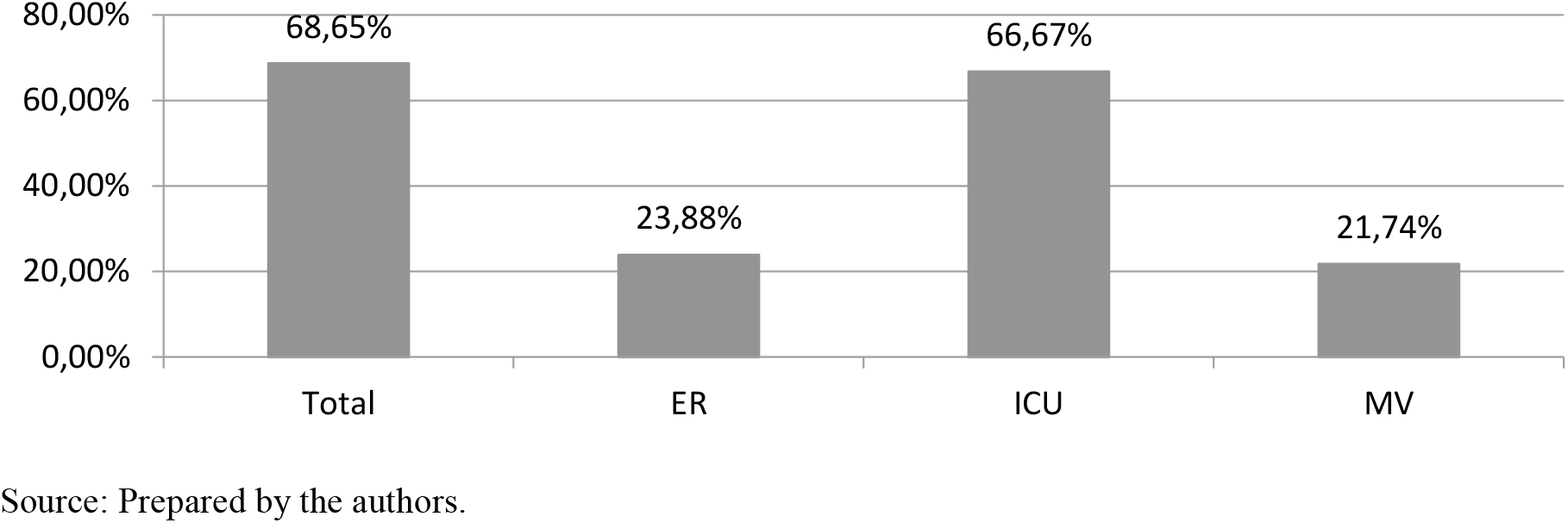
Mortality rate by type of treatment

**Graph 3.**
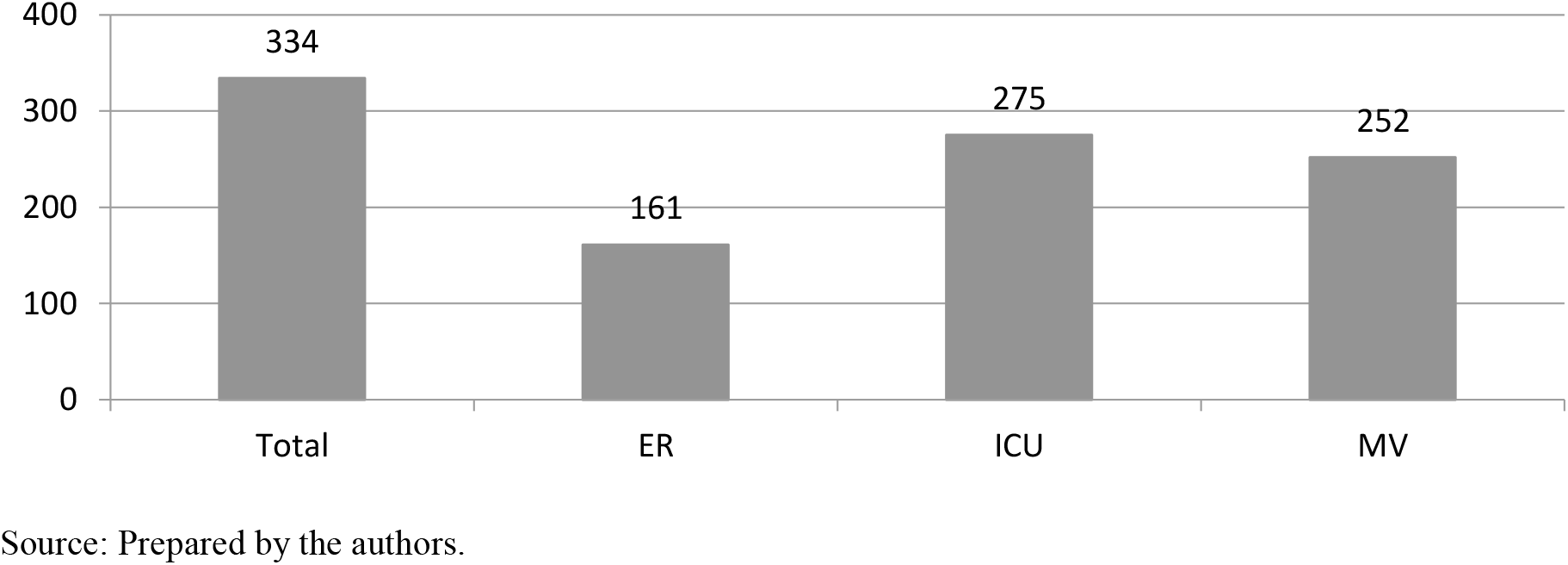
Length of stay in hours by type of treatment

The total mortality rate of the sample, which comprises the period in which the patient entered the ER and / or was intubated, until the end of the treatment (discharge or death), was 68.65%. Of the 67 treated patients, 46 patients died, 20 patients were discharged and one patient was still on treatment. The average hospital stay was 334: 36h (13 days, 22 hours and 36 minutes).

Of the 67 patients admitted to the ER, 45 were transferred to the ICU and 22 were treated at this unit. Of the patients who continued on treatment at ER 06, they were discharged and 16 died. The average hospital stay was 161: 52h (06 days, 17 hours and 52 minutes) and the Mortality Rate was 23.88%. 45 patients were admitted to the ICU, of these 30 died, 14 were discharged and one continued on treatment. The average hospital stay was 275: 14h (11 days, 11 hours and 14 minutes) and the Mortality Rate was 66.67%.

Of the intubated patients (n = 67), only 23 patients were extubated, 41 died during treatment with MV and 03 patients did not have the information on the date and time of extubation. However, of these 03 patients without information, 02 were discharged and 01 died. For patients who were intubated and extubated (n = 23) 18 survived and were discharged and 05 died after extubation, the survival rate after extubation was 78.26% and the mean MV time was 252: 15h (10 days, 12 hours and 15 minutes). Analyzing the total sample (n = 67), the general ratio was for every 03 patients who entered for treatment, 01 was discharged and 02 died. Considering only the patients who were under treatment at the ER (n = 22), it was found that, for every 03 patients who were hospitalized at the ER, 02 patients were discharged and 01 patient died before being admitted to the ICU. In the ICU (n = 45) for every 04 inpatients, 02 were discharged and 02 died. And, of the patients who received treatment with MV (n = 23), for every 04 patients, 03 were discharged and 01 died.

It was observed that patients who spent less time in treatment were more likely to recover and be discharged, as seen in the mean time of treatment in ER and MV. It was also found that patients who received treatment with MV had a better result, since the largest proportion of intubated and extubated patients were discharged.

In this analyzed sample, these were the results found. However, it is essential to point out that the final outcome of each treated patient is unique, since each individual reacts in a specific way to each treatment performed and there are patients who recover with more or less hospitalization time. In this sense, for a more in-depth analysis, it would be necessary to incorporate other information about the care protocol adopted in each case and thereby verify the variables that interfere in the patient’s recovery beyond the treatment time.

## DISCUSSION

In this study, it is possible to evidence a significant sample of patients. Therefore, this hospital accompanies patients in ER and ICU for a long period, therefore, prolonged hospitalization is associated with longer hospital stay and is conducive to increased mortality. It is observed that the shorter the treatment time, the greater the possibility of recovery and discharge. However, compared to the needs of patients, the mortality rate obtained has become high.

Through the survey carried out for a period of 4 months, the average length of stay was 334: 36h, with a mortality rate of 68.65%. According to the Society of critical care medicine, a critical patient must be transferred from the emergency room to the ICU within a maximum of 6 hours. In a prospective cohort study, which took place from January to December 2014; 6,176 patients were treated in the emergency room, among whom 1,913 (31%) needed a bed in the ICU. The median time spent in the emergency room was 17 hours (9 – 33 hours). Hospitalization for infection / sepsis was an independent predictor for prolonged length of stay in the unit (RC: 2.75; 95% CI 1.38 – 5.48, p = 0.004), but the waiting time for admission to the ICU was not. The mortality rate was higher in Group 3 (38%) than in Group 1 (31%), but the difference was not statistically significant31. Therefore, the results of our research corroborate the research of SANTOS, MACHADO & LOBO (2020)^31^.

It was observed that patients who had spent less time in both the ER and the ICU had a greater probability of recovering and being discharged. In another study of a non-randomized clinical trial analyzed about of 466 patients in relation to the time of IMV and length of stay in the ICU in three ICUs of a hospital in Caxias do Sul (RS). Where 235 and 231 patients were evaluated in the pre-intervention and post-intervention phases, respectively. After the implementation of the checklist, there were significant reductions in the medians (interquartile ranges) of the length of stay in the ICU – from 8 (4–17) days to 5 (3–11) days; p ≤ 0.001 – and in the IMV time – from 5 (1–12) days to 2 (< 1–7) days; p ≤ 0.001. This study concluded that the application of routine multi-professional checklists is related to the reduction in the number of days of IMV and ICU stay^32^. However, the results of this study are related to the results of this research.

According to Santos (2019) who carried out an exploratory descriptive study in the state of Rio de Janeiro, it was possible to identify that, although the number of beds was reduced in the analyzed period, the number of hospitalizations increased significantly. There was also an increase in the number of emergency admissions, while elective admissions decreased. Analyzing the relationship between the number of possible admissions and the number of hospitalizations performed, we could observe a significant improvement through the available beds^33^.

It was also observed that even with almost 60% of the city’s territory covered by the Family Health Strategy, there are still many hospitalizations sensitive to primary care, it has also been shown that this type of hospitalization is falling. The study made it possible to know that the reduction in hospitalization of public health units, may mean a threat to the principles of rights guaranteed by the 1988 federal constitution. In this scenario, in which there may be compromise in the SUS service line, assess whether there was a drop in the number of hospitalizations can be a warning sign of great importance for health managers^33^. The outcomes of this prospective cohort study were sanctioned with the results of our research.

A total of 6176 patients were admitted to the emergency room, of whom 1913 (31%) had an ICU vacancy. A total of 209 patients (11%) from the emergency room were admitted to the mixed ICU of this hospital. The median time spent in the emergency room was 17 hours [9 to 33 hours]. Patients admitted to the ICU more quickly were younger (group 1, 48 years (median), 28–61 years [25% –75%], group 2, 52 years [31–60 years], group 3, 58 years [44 –72 years]; (p = 0.001). The length of hospital stay increased significantly in group 3 (18 days, [9–31 days]) compared with group 1 (10 days, [4–21 days]) and group 2 (11 days, [6.5–20 days] (p = 0.002)). The mortality rate increased from 31% in the first group to 38% in the third group (p = 0.639). With the following results they concluded that the increase in waiting time for admission to the ICU was associated with prolonged hospital stay and increased use of resources^34^.

## CONCLUSION

Based on the analyzes carried out in this research, we concluded that the length of stay in the ER and in the ICU, correlated to patients on ventilatory support, is related to the mortality rate.

Due to overcrowding giving priority to the hospital sectors (ER and ICU) in recent years, there is a cordial rise in the number of beds to meet the necessary demand of individuals who lack this type of service. Prolonged hospitalization is associated with longer hospital stays and is conducive to increased mortality. With a view to better regulation, it is essential to redefine and consolidate the central regulation as an essential policy to manage the demands of critically ill patients and make available resources available at the right time. Therefore, the National Regulatory Center can work together with all stakeholders to ensure clinical governance and consolidate the organization of the network.

Care in the hospital environment in the ER sector is differentiated from that of the ICU, which is managed by a specialized multidisciplinary team, always aiming at the best care for critically ill patients, and constantly improving to maintain excellence in advances in intensive care to Cheers. While in the ER there is a specialized multidisciplinary team, but which manages several sectors. Thus, we conclude that the care in the ER is definitely not adequate when compared to the care in the ICU, in view of an environment with less restrictions, and in the ICU obtaining greater human resources.

Therefore, by increasing the demand for beds, we will reduce the waiting list for critical patients who need advanced intensive services, thus, providing the best necessary care, we can reduce mortality rates, aimed at waiting for ICU vacancies.

## Data Availability

All data availability inn Brazilian Department of databases - www.datasus.gov.br

